# Low-attenuation Coronary Plaque Volume and Cardiovascular Events in Patients with Distinct Metabolic Phenotypes

**DOI:** 10.1101/2023.03.02.23286724

**Authors:** Kenichiro Otsuka, Hirotoshi Ishikawa, Kenei Shimada, Kana Hojo, Hiroki Yamaura, Yasushi Kono, Noriaki Kasayuki, Daiju Fukuda

## Abstract

**Background:** This study aimed to investigate the association between diabetes mellitus (DM), high-risk coronary plaque burden, and risk of cardiovascular outcomes across metabolic phenotypes in patients with suspected coronary artery disease (CAD) who underwent coronary computed tomography angiography (CCTA).

**Methods:** We included 530 patients who underwent CCTA. Metabolic syndrome (MetS) was defined as the presence of a visceral adipose tissue area ≥ 100 cm^2^ in patients with DM (n = 58), or two or more MetS components excluding DM (n = 114). Remaining patients were categorized into non-MetS patients with DM (n = 52) and non-MetS patients without DM (n = 306). CCTA-based high-risk plaque was defined as low-attenuation plaque (LAP) volume > 4 %. Primary endpoint was presence of a major cardiovascular event (MACE), which was defined as a composite of cardiovascular death, acute coronary syndrome, and coronary revascularization.

**Results:** Incidence of MACE was highest in the non-MetS with DM group, followed hierarchically by the MetS with DM, MetS without DM, and non-MetS without DM groups. In the multivariable Cox hazard model analysis, DM as a predictor was associated with MACE independent of LAP volume > 4 % (hazard ratio, 2.68; 95% confidence interval, 1.16–6.18; p = 0.02), although MetS did not remain an independent predictor. LAP volume > 4 % remained a predictor of MACE independent of each metabolic phenotype or DM.

**Conclusions:** This study demonstrated that DM, rather than MetS, is a predictor of coronary events independent of high-risk plaque volume in patients who underwent CCTA.

**Clinical Perspective:**

1. What Is New?

- This study investigated the association between diabetes mellitus (DM), high-risk coronary plaque burden, and major adverse cardiovascular events (MACE) across metabolic phenotypes stratified by the presence or absence of metabolic syndrome (MetS) and DM in patients with suspected coronary artery disease (CAD) who underwent coronary computed tomography angiography (CCTA).
- Among the four metabolic phenotypes, incidence of MACE was highest in the non-MetS with DM group, followed hierarchically by the MetS with DM, MetS without DM, and non-MetS without DM groups. Low-attenuation coronary plaque (LAP) volume > 4% was a robust predictor of MACE among the metabolic phenotypes. Furthermore, DM, independent of LAP volume > 4%, was a predictor of MACE, while MetS did not show a significant predictive value.
2. What Are the Clinical Implications?

- Our results demonstrate that individuals with DM alone have a significantly higher risk of developing cardiovascular events than those with MetS, indicating that DM is an independent predictor of cardiovascular events irrespective of the presence of obstructive CAD or LAP volume greater than 4%.

## Introduction

The global prevalence of diabetes mellitus (DM) and metabolic syndrome (MetS) has increased significantly over the past decades, contributing to an increased risk of atherosclerotic cardiovascular disease (ASCVD) ^1^. Clinical studies have demonstrated that an overweight status (25–29.9 kg/m^2^) or obesity (≥ 30 kg/m^2^), defined by body mass index (BMI) alone, reflects heterogeneous body fat distribution and distinct metabolic conditions ^2^. This raises questions about the relationship between BMI and ASCVD risk, which leads to the obesity paradox. Although visceral adiposity, a modifiable risk factor for MetS, helps identify metabolically unhealthy individuals ^1–4^, coronary plaque features associated with ASCVD events in individuals with distinct metabolic phenotypes remain largely unknown.

Coronary computed tomography angiography (CCTA) facilitates the diagnosis of coronary artery disease (CAD) and offers a prognostic value based on high-risk coronary plaque features beyond stenosis severity ^5^. Furthermore, a recent CCTA study demonstrated that low-attenuation non-calcified coronary plaque burden is the strongest prognostic marker among other clinical factors, such as the presence of CAD and coronary artery calcium score (CACS) ^6^. This finding suggests the utility of CCTA in identifying high-risk patients. In a sub-analysis of a large clinical CCTA trial of patients with chest pain and distinct metabolic phenotypes, Kammerlander et al. demonstrated that metabolically unhealthy individuals without obesity were at a high risk for ASCVD events^7^. Although DM plays a key role in the pathophysiology of MetS ^8^, there is limited knowledge on its association with high-risk plaque volume detected on CCTA and consequent cardiovascular outcomes. In this CCTA study, we aimed to investigate the association between MetS with and without DM, imaging findings, and the risk of outcomes across MetS phenotypes.

## Methods

The data that support the findings of this study are available from the corresponding author on reasonable request.

### Study population and metabolic phenotypes

This study included patients who underwent CCTA between January 2018 and December 2020 and were clinically indicated for stable chest symptoms, along with an abdominal computed tomography (CT) scan to assess abdominal obesity. The exclusion criteria were as follows: (1) acute coronary syndrome; (2) previous history of coronary artery bypass graft or open-heart surgery; (3) congestive heart failure; (4) previous history of percutaneous coronary intervention; (5) insufficient patient information; and (6) loss to follow-up. Figure 1 illustrates the flowchart of the study population comprising 530 patients. All participants provided written informed consent for the use of de-identified data, including clinical information, laboratory tests, and CCTA imaging. The Fujiikai Kashibaseiki Hospital Institutional Review Board approved pooled data analysis (2021-A).

**Figure 1.**
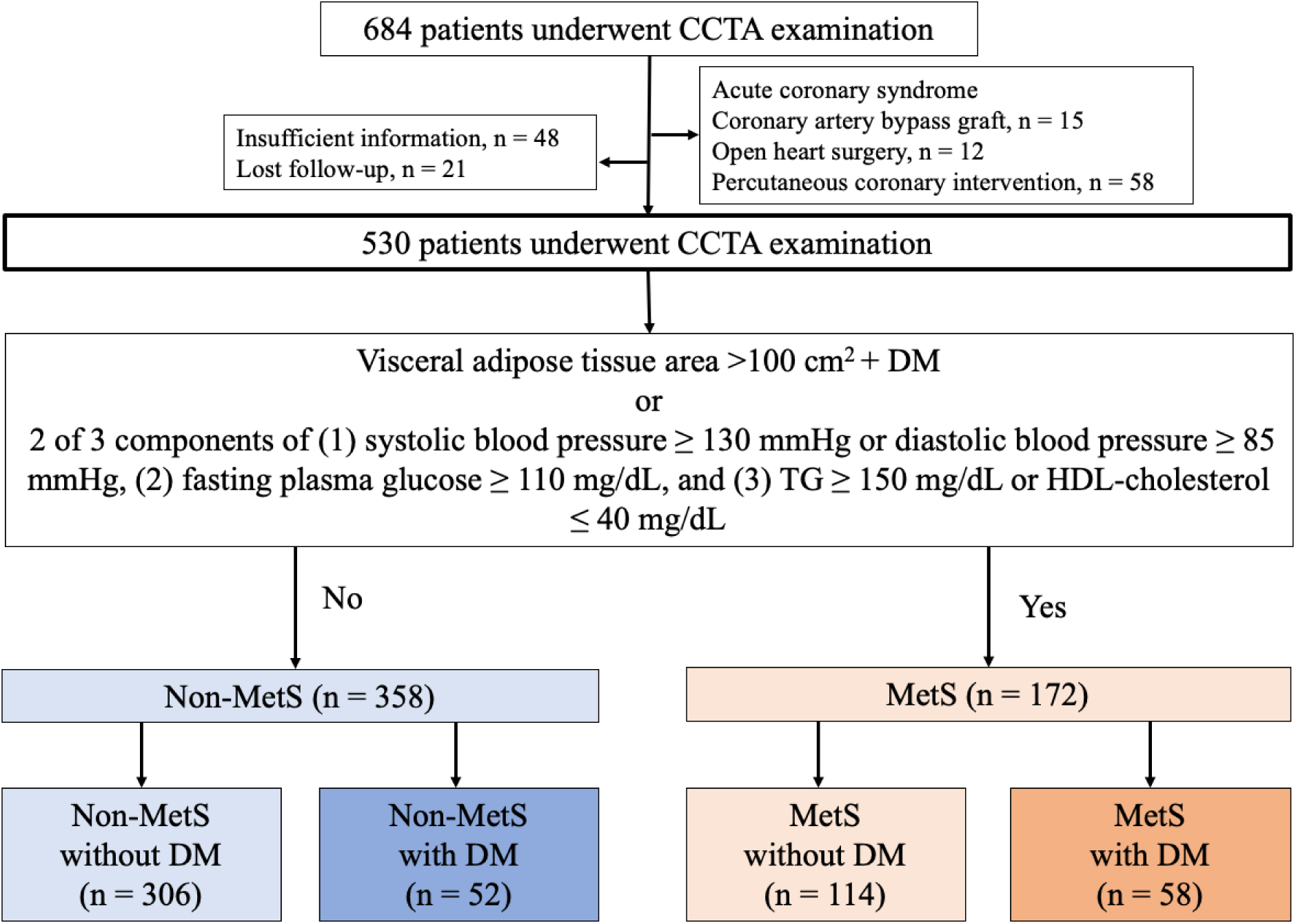
Flow chart of study patients. A flowchart depicting the 530 patients who underwent CCTA examination and how they were divided into No-MetS and MetS groups. Patients were further categorized into four groups according to the presence or absence of metabolic equivalents (MetS) and DM. CCTA, coronary computed tomography angiography; DM, diabetes mellitus; MetS, metabolic syndrome.

Non-contrast-enhanced abdominal CT imaging was performed before CCTA to measure the visceral adipose tissue (VAT) areas. VAT areas were measured at the L2–L3 level using a dedicated software (SYNAPSE VINCENT, Fujifilm Inc., Tokyo, Japan). In the present study, abdominal obesity was defined as a VAT area ≥100 cm^2^, corresponding to an abdominal circumference ≥ 85 cm for men and ≥ 90 cm for women ^9^. MetS was defined based on the Japanese Committee for the Diagnostic Criteria of Metabolic Syndrome ^10^. MetS was defined as the presence of abdominal obesity with DM (MetS with DM) or two or more of the following components in the absence of DM: (1) systolic blood pressure (BP) ≥130 mmHg or diastolic BP ≥85 mmHg; (2) fasting plasma glucose ≥110 mg/dL; and (3) triglycerides ≥150 mg/dL or high-density lipoprotein cholesterol ≤40 mg/dL. In addition, medical therapy using antihypertensive, antidiabetic, and lipid-lowering drugs was considered an MetS component. Patients were further categorized into four groups according to the presence or absence of metabolic equivalents (MetS) and DM. The triglyceride-glucose (TyG) index was used to estimate the insulin resistance in the four groups ^11^.

Pharmacological treatment and lifestyle modifications were recommended for all patients according to the guidelines for hypertension, dyslipidemia, and diabetes, if present ^12^. In addition, patients underwent invasive coronary angiography or coronary revascularization based on the results of the CCTA and noninvasive stress tests ^13^. According to the Japanese Atherothrombosis Society guidelines, the treatment targets of low-density lipoprotein cholesterol (LDL-C) for the primary prevention of ASCVD were <100 mg/dL for high-risk, 120 mg/dL for moderate-risk, or 140 mg/dL for low-risk patients ^12^. The treatment target for the fasting triglyceride (TG) level was < 150 mg/dL. For hypertensive patients, the treatment target for blood pressure is <140/90 mmHg, particularly for patients with diabetes or CKD with albuminuria <130/80 mmHg ^14^. The target glycemic control was HbA1c < 7.0% for preventing diabetic complications and < 8.0% for those with difficulties in glycemic control for hypoglycemia ^15^.

### CCTA imaging and analysis

CCTA was performed using a 320-row multi-detector CT scanner (Aquilion ONE/NATURE Edition, Canon Medical Systems, Inc., Japan) ^16^. The scan parameters included a detector collimation of 0.5 × 320 mm, gantry rotation time of 350 ms, tube voltage of 120 kV, and tube current of 130–600 mA. An electrocardiogram-triggered prospective gating method was used for CCTA. CACS was evaluated using the Agatston method at a fixed thickness of 3 mm ^17^. Images were reconstructed using forward projected model-based iterative reconstruction solution for coronary artery analysis, with a cross-sectional thickness of 0.5 mm and a reconstruction increment of 0.25 mm.

The Agatston score categories were as follows: 0, 1–100, 101–400, and >400 Agatston units, using a software (SYNAPSE VINCENT version 4.6., Fujifilm inc., Tokyo, Japan). Coronary artery diameter stenosis was reported based on a 16-segment American Heart Association model by two observers (K.O. and H.I.). Obstructive CAD was defined as the presence of coronary plaques with diameter stenosis of ≥ 50% in one or more major epicardial vessels, and/or ≥ 50% in the left main coronary segment. Non-obstructive CAD was defined as having less than 50% diameter stenosis of epicardial coronary arteries. Patients who did not fall in either categories were categorized as not having CAD.

For coronary plaque analysis, coronary artery centerlines were identified semi-automatically; the proximal and distal portions of the coronary plaque lesions were manually defined; and the vessel wall, lumen, and plaque components were auto-segmented and manually adjusted. Lesions were categorized based on their composition into calcified plaques (CP) (> 150 Hounsfield units [HU]) and non-CP (NCP) (< 150 HU). Furthermore, a low-attenuation plaque (LAP) was defined as a region with a CT value < 30 HU ^16^. The percentage of plaque volume for each component was calculated as the plaque volume divided by the vessel volume.

The epicardial adipose tissue (EAT) volume was measured from contrast-enhanced CT images using the VINCENT software, cited previously ^16, 18^. Several equidistant axial planes were extracted based on the heart size. The upper limit of the slice was set at the bifurcation of the pulmonary artery trunk, whereas the lower limit was set at the last slice containing any structure of the heart. In each plane, the software autodetected a smooth, closed pericardial contour as the region of interest; adipose tissue was identified with CT attenuation values ranging from -250 to -30 HU within the pericardial sac ^16, 18^. Finally, EAT volume was calculated as the sum of the EAT areas in each slice. The mean CT value within the measured EAT volume was reported.

### Endpoints

The primary endpoint was major adverse cardiovascular events (MACE) defined by a composite of cardiovascular death, nonfatal myocardial infarction, unstable angina, or symptom- or ischemia-driven coronary revascularization. Cardiovascular death was defined as death due to cardiovascular causes including myocardial infarction, sudden cardiac arrest, heart failure, and stroke ^19^. Non-fatal myocardial infarction was defined as typical persistent chest pain with elevated cardiac enzymes ^19^. Unstable angina was defined as new-onset angina, exacerbation of angina with light exertion, or angina at rest without elevated cardiac enzyme levels. Symptom- or ischemia-driven coronary revascularization was defined as coronary revascularization >3 months after CCTA imaging at baseline, with >75% diameter stenosis with symptoms, or positive functional tests, or with a ≥90% diameter stenosis with symptoms.

### Statistical analysis

All statistical analyses were performed using SPSS version 24 software (IBM Corp., Armonk, NY, USA). Categorical variables were presented as absolute and relative frequencies, and continuous variables were presented as mean (±standard deviation). Subject characteristics were compared using one-way analysis of variance for numerical variables. Categorical variables were analyzed using the chi-square or Fisher’s exact tests. Multivariate Cox proportional hazards analysis was used to estimate hazard ratios (HRs) with 95% confidence intervals (CIs). To test the hypothesis that DM or MetS serve as a predictor of MACE independent of high-risk plaque volume, DM (or MetS) and LAP volume > 4% were entered into the multivariate Cox proportional hazards models adjusted for the Suita CVD risk score (model 1 and model 2). Furthermore, to test the hypothesis that each metabolic phenotype remains as a predictor of MACE independent of high-risk plaque volume, each metabolic phenotype, as compared with non-DM without MetS and LAP volume > 4%, were entered into the multivariate Cox proportional hazards models adjusted for the Suita CVD risk score (models 3, 4, and 5).

Additionally, multivariable models were constructed. Variables entered into the multivariate models were selected on the basis of clinical plausibility, multicollinearity, and known association with CV death and acute coronary syndrome (ACS), with the adjustment for Suita CVD risk score: Model 1, LAP volume > 4% and TyG index; Model 2, LAP volume > 4% and CKD; Model 3, LAP volume > 4% and CRP; and Model 4, LAP volume > 4% and obstructive CAD (any vessels) (supplementary Table 1).

Kaplan–Meier curves and log-rank tests were used to depict and assess the differences in cumulative event rates among the four groups. Analyses were initiated at the time of CCTA imaging and terminated at the earliest occurrence of the primary endpoint or at the median follow-up (2.91 years). Analyses were censored at the last follow-up or at a composite event, whichever occurred earlier. Statistical significance was set at p < 0.05 (two-sided).

## Results

All CCTA images of the 530 patients qualified for analysis. The mean age of the patients was 64 ± 14 years, and 299 (56%) patients were men. Baseline characteristics stratified by metabolic phenotype are presented in Table 1. MetS were present in 172 of the 530 patients (32%), and DM was observed in 110 (21%). Of these, DM was present in 52 patients in the non-MetS (15%) and 58 in the MetS group (44%). The highest TyG index was observed in the patients with metabolic equivalents (MetS) and DM, followed by those without DM, non-MetS with DM, and non-MetS without DM.

**Table 1.**
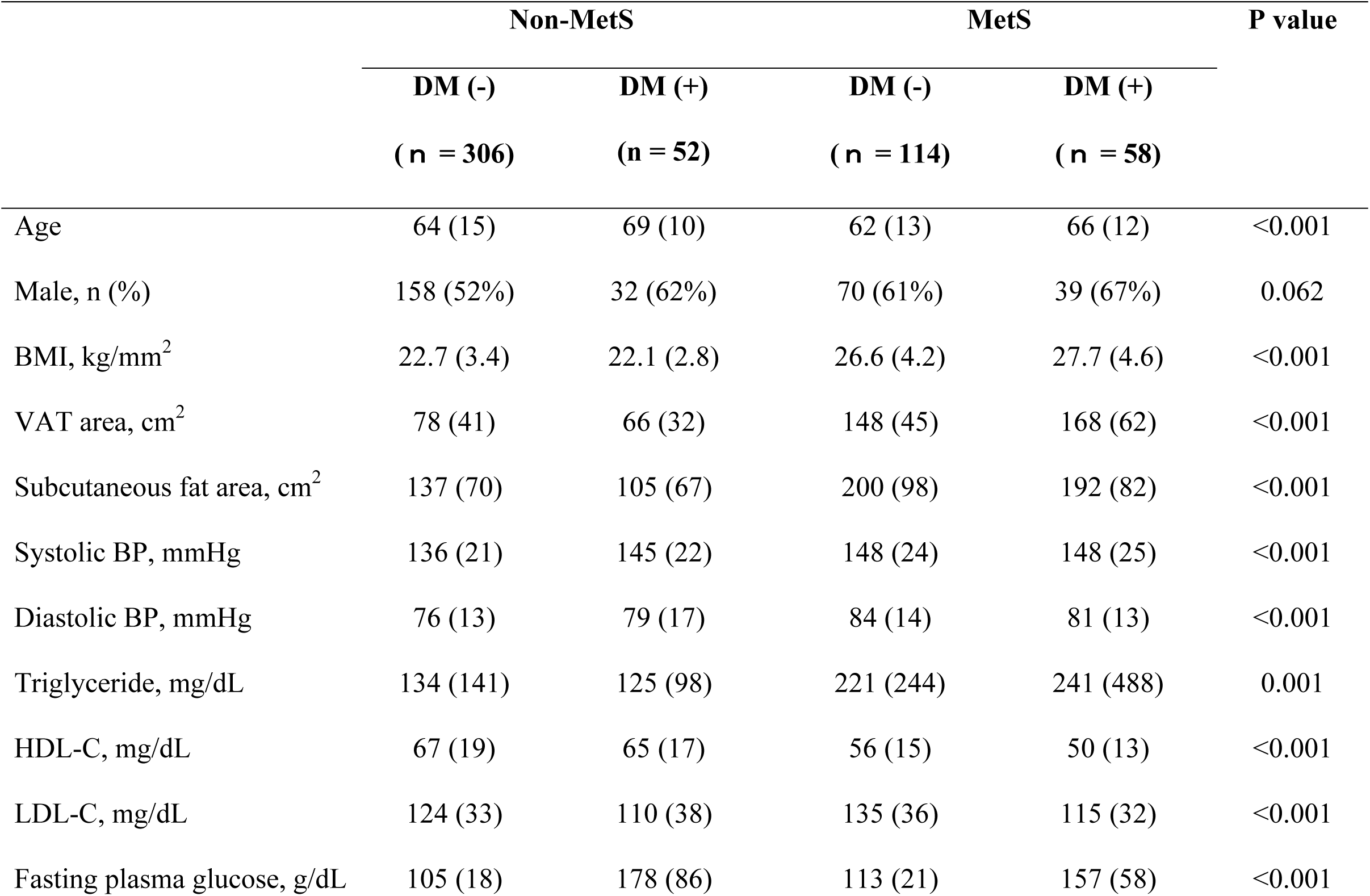
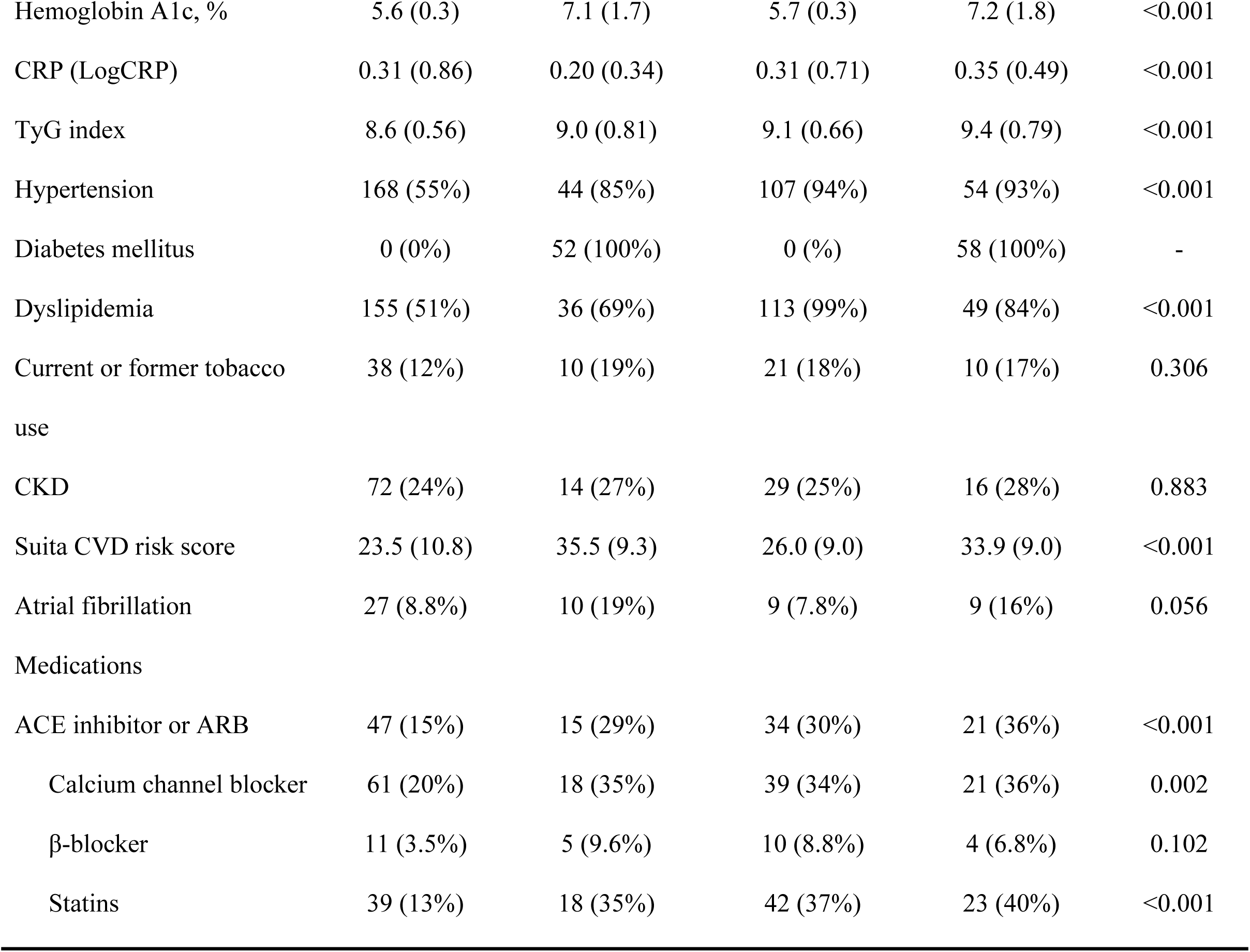
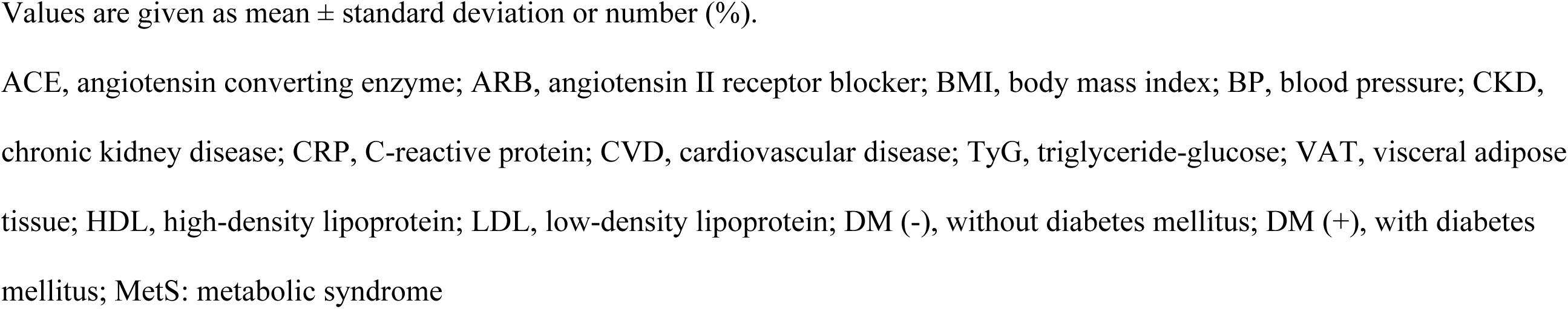
Patient characteristics according to metabolic phenotypes

Table 2 shows the baseline CCTA findings of the four groups. The prevalence of CACS > 400 was greatest in the MetS with DM (22%) and non-MetS with DM groups (21%, overall p < 0.001). The frequency of obstructive CAD was the highest in the non-MetS with DM group (58%), sequentially followed by the MetS with DM (46%), MetS without DM (39%), and non-MetS without DM groups (36%). The MetS with DM group featured greater NCP, LAP, and CP volumes than those without DM. While significantly greater EAT volumes were observed in individuals with MetS than in those without MetS, there was no significant difference in DM. Non-MetS with DM had the lowest mean CT values among the four groups (p < 0.001).

**Table 2.**
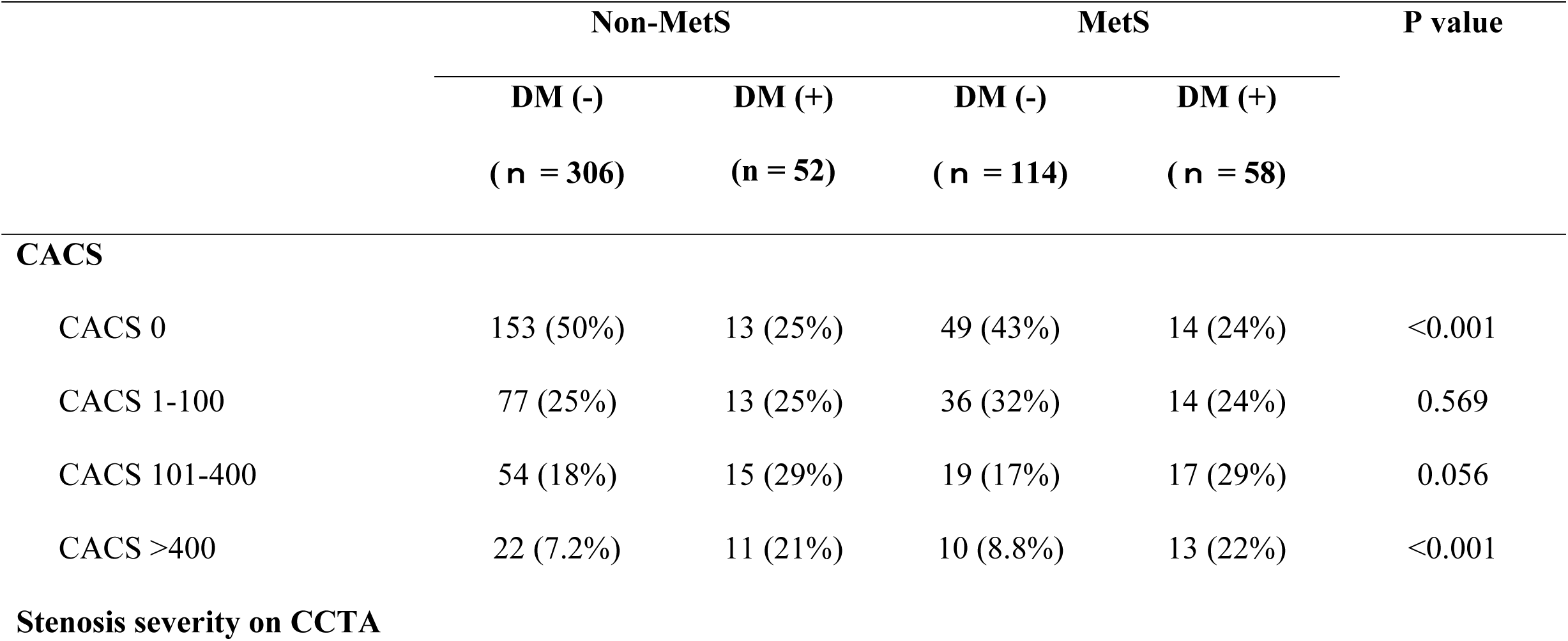
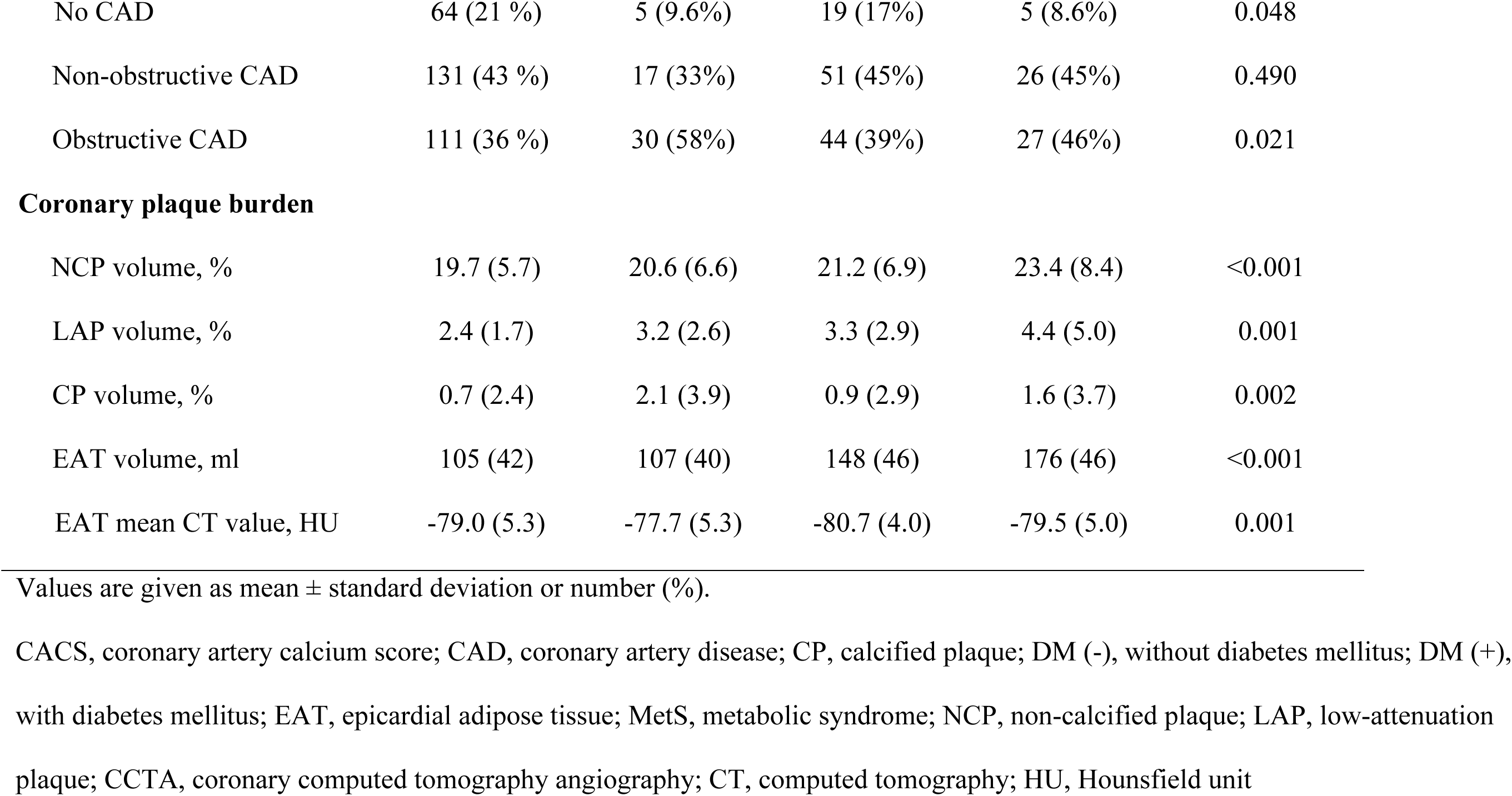
CCTA findings according to metabolic phenotypes

|  | Non-MetS |  | MetS |  | P value |
| --- | --- | --- | --- | --- | --- |
|  | DM (-) | DM (+) | DM (-) | DM (+) |  |
|  | (n = 306) | (n = 52) | (n = 114) | (n = 58) |  |
| **CACS** |  |  |  |  |  |
| CACS 0 | 153 (50%) | 13 (25%) | 49 (43%) | 14 (24%) | <0.001 |
| CACS 1-100 | 77 (25%) | 13 (25%) | 36 (32%) | 14 (24%) | 0.569 |
| CACS 101-400 | 54 (18%) | 15 (29%) | 19 (17%) | 17 (29%) | 0.056 |
| CACS >400 | 22 (7.2%) | 11 (21%) | 10 (8.8%) | 13 (22%) | <0.001 |
| **Stenosis severity on CCTA** |  |  |  |  |  |
| No CAD | 64 (21 %) | 5 (9.6%) | 19 (17%) | 5 (8.6%) | 0.048 |
| Non-obstructive CAD | 131 (43 %) | 17 (33%) | 51 (45%) | 26 (45%) | 0.490 |
| Obstructive CAD | 111 (36 %) | 30 (58%) | 44 (39%) | 27 (46%) | 0.021 |
| **Coronary plaque burden** |  |  |  |  |  |
| NCP volume, % | 19.7 (5.7) | 20.6 (6.6) | 21.2 (6.9) | 23.4 (8.4) | <0.001 |
| LAP volume, % | 2.4 (1.7) | 3.2 (2.6) | 3.3 (2.9) | 4.4 (5.0) | 0.001 |
| CP volume, % | 0.7 (2.4) | 2.1 (3.9) | 0.9 (2.9) | 1.6 (3.7) | 0.002 |
| EAT volume, ml | 105 (42) | 107 (40) | 148 (46) | 176 (46) | <0.001 |
| EAT mean CT value, HU | -79.0 (5.3) | -77.7 (5.3) | -80.7 (4.0) | -79.5 (5.0) | 0.001 |

Values are given as mean ± standard deviation or number (%).
CACS, coronary artery calcium score; CAD, coronary artery disease; CP, calcified plaque; DM (-), without diabetes mellitus; DM (+), with diabetes mellitus; EAT, epicardial adipose tissue; MetS, metabolic syndrome; NCP, non-calcified plaque; LAP, low-attenuation plaque; CCTA, coronary computed tomography angiography; CT, computed tomography; HU, Hounsfield unit

### Primary outcome

During a mean follow-up period of 2.7 ± 0.9 years (median 2.91 years), MACE was observed in 25 patients (4.7%). The primary outcome was observed in seven patients in the non-MetS without DM group, eight in the non-MetS with DM group, six in the MetS without DM group, and four in the MetS with DM group. The incidence rate for the composite endpoint was highest in the non-MetS with DM group, followed hierarchically by the MetS with DM, MetS without DM, and non-MetS without DM group (p < 0.001, log-rank test; Figure 2).

**Figure 2.**
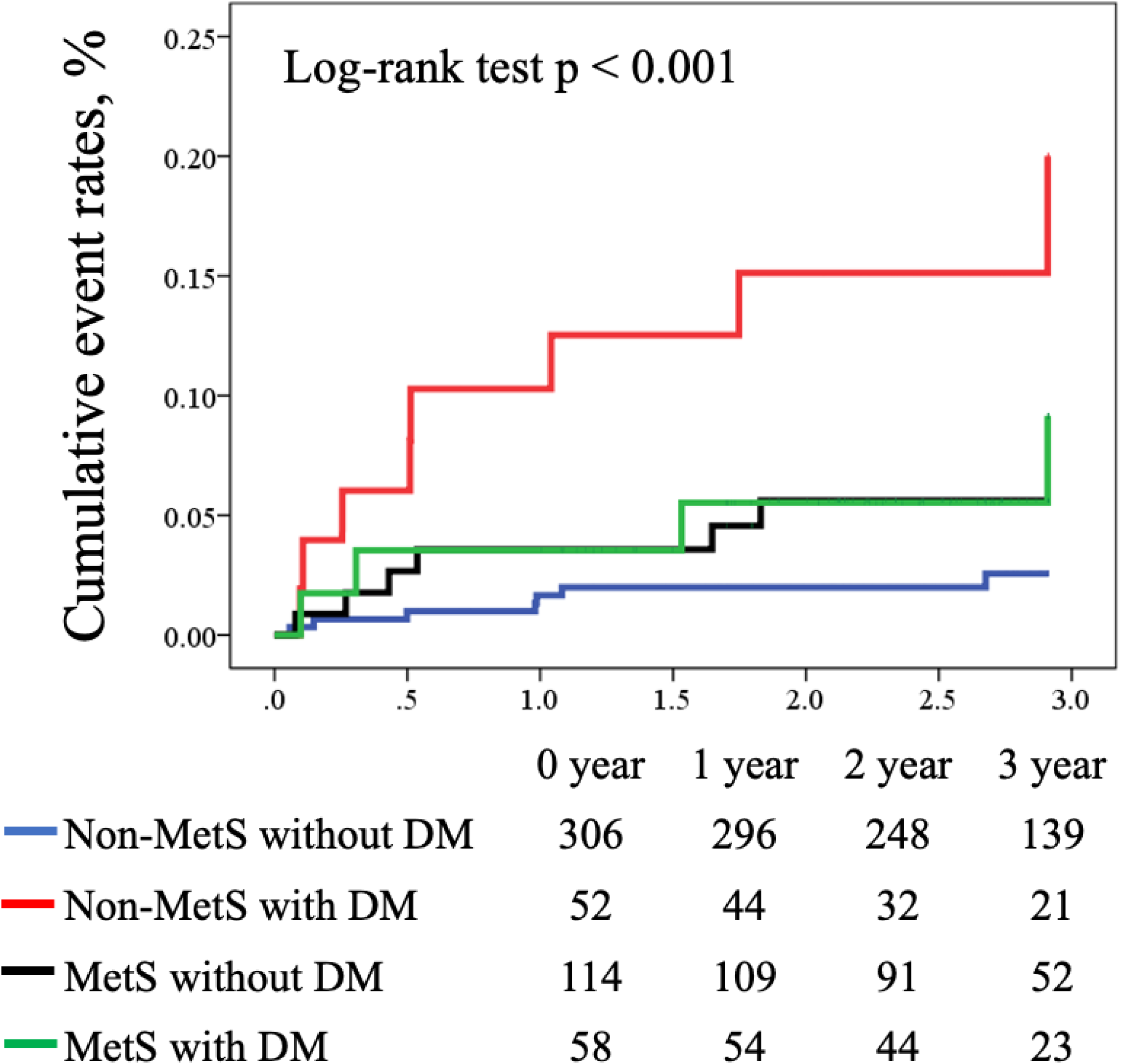
Kaplan–Meier analysis for prediction of MACE stratified by metabolic phenotypes. Kaplan–Meier curves demonstrating significant differences between metabolic phenotypes in cumulative event rates, using a composite endpoint of cardiovascular death, acute coronary syndrome, and symptom- or ischemia-driven coronary revascularization. Higher event rates were present in non-MetS patients with DM (log-rank test, p <0.001), followed by MetS with DM (log-rank test, p <0.001). DM, diabetes mellitus; MetS, metabolic syndrome; MACE, major adverse cardiovascular event.

Table 3 summarizes the unadjusted Cox proportional hazards models used to predict the primary endpoint. CACS > 400 (p < 0.001), LAP volume > 4% (p < 0.001), obstructive CAD (patient-level and vessel-level, p < 0.05), and DM (p < 0.001) were significantly associated with the primary endpoint. Patients with angiographically confirmed MACE (n = 20) exhibited significantly higher prevalence of LAP volume > 4% in LAD (45% versus 23.5%; p = 0.036) and RCA (40% versus 18%; p = 0.035) and tended to have higher prevalence of LAP volume > 4% in LCX (20% vs. 9.3%; p = 0.09) compared to those without MACE. In the multivariable Cox proportional hazards model analysis (Table 4), DM as a predictor was associated with the primary endpoint, independent of LAP volume > 4% (HR for DM in model 1; HR, 2.68; 95% CI, 1.16–6.18; p = 0.02), although MetS did not remain an independent predictor (model 2 in Table 4). For the subgroup analysis of each metabolic phenotype (non-DM without MetS as reference), DM without MetS remained as a predictor of MACE independent of LAP volume > 4% (model 3), whereas DM with MetS (model 4) or non-DM with MetS (model 5) did not reach statistical significance. Finally, we examined the association of MACEs with parameters other than DM or MetS. In the multivariable models, TyG index (HR, 1.78; 95% CI, 1.10–2.89; p = 0.019) or obstructive CAD (HR, 5.49; 95% CI, 1.84–16.38; p = 0.002) was independently associated with MACE even after adjusting for the Suita CVD risk score, but CKD or CRP did not remain as a predictor (supplementary Table 1).

**Table 3.**
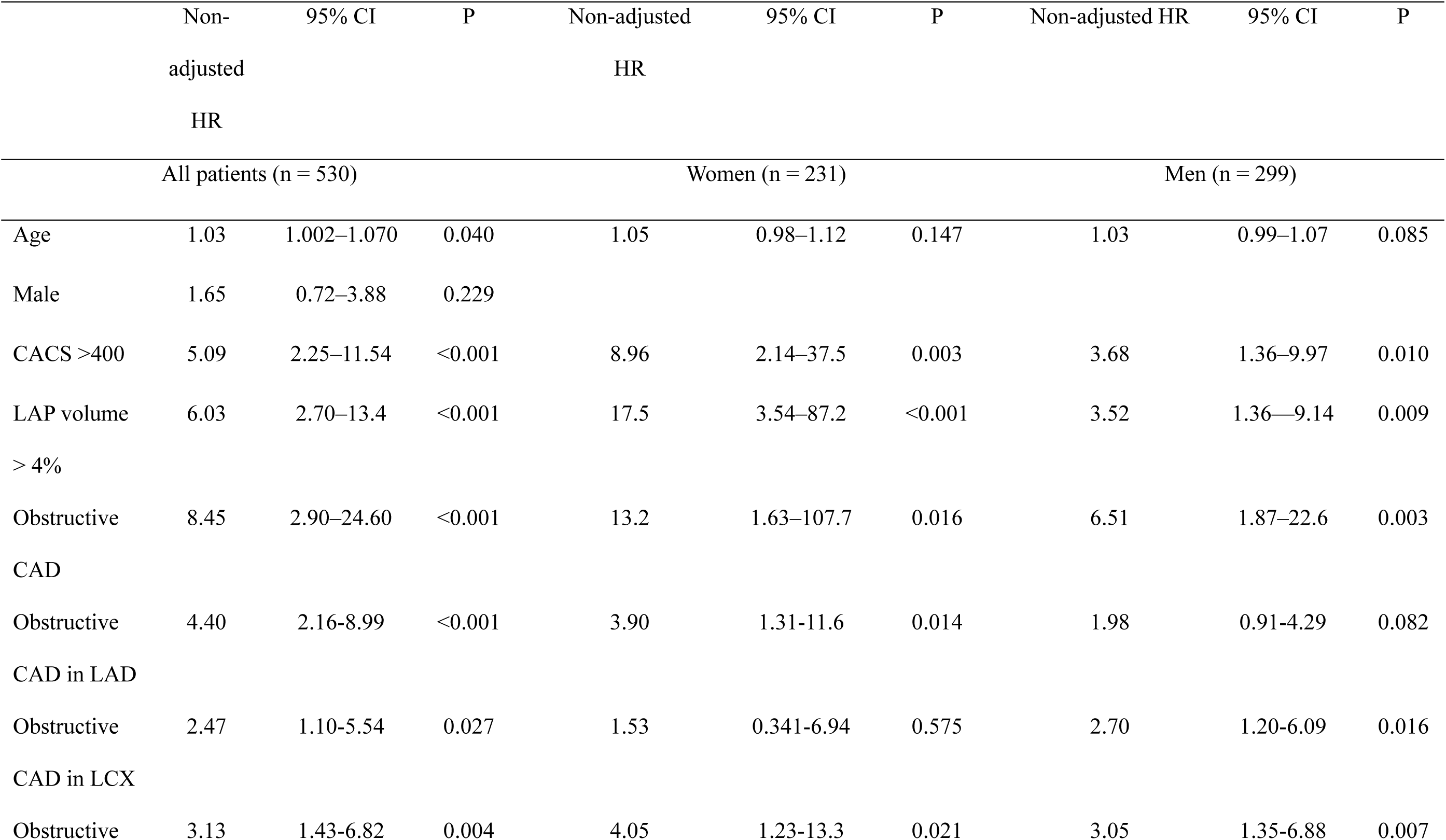
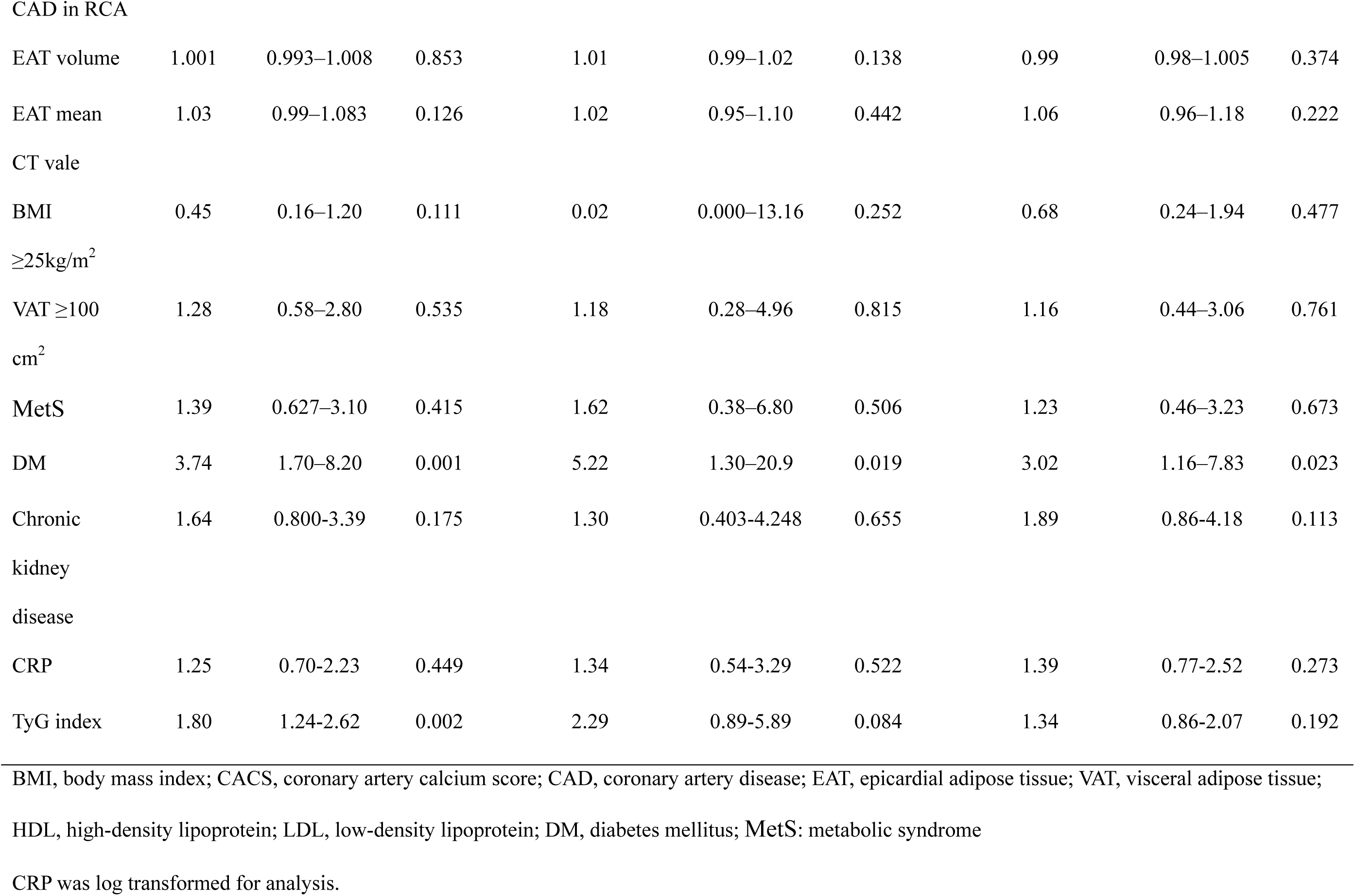
Unadjusted Cox proportional hazards model to predict the primary outcome

**Table 4.**
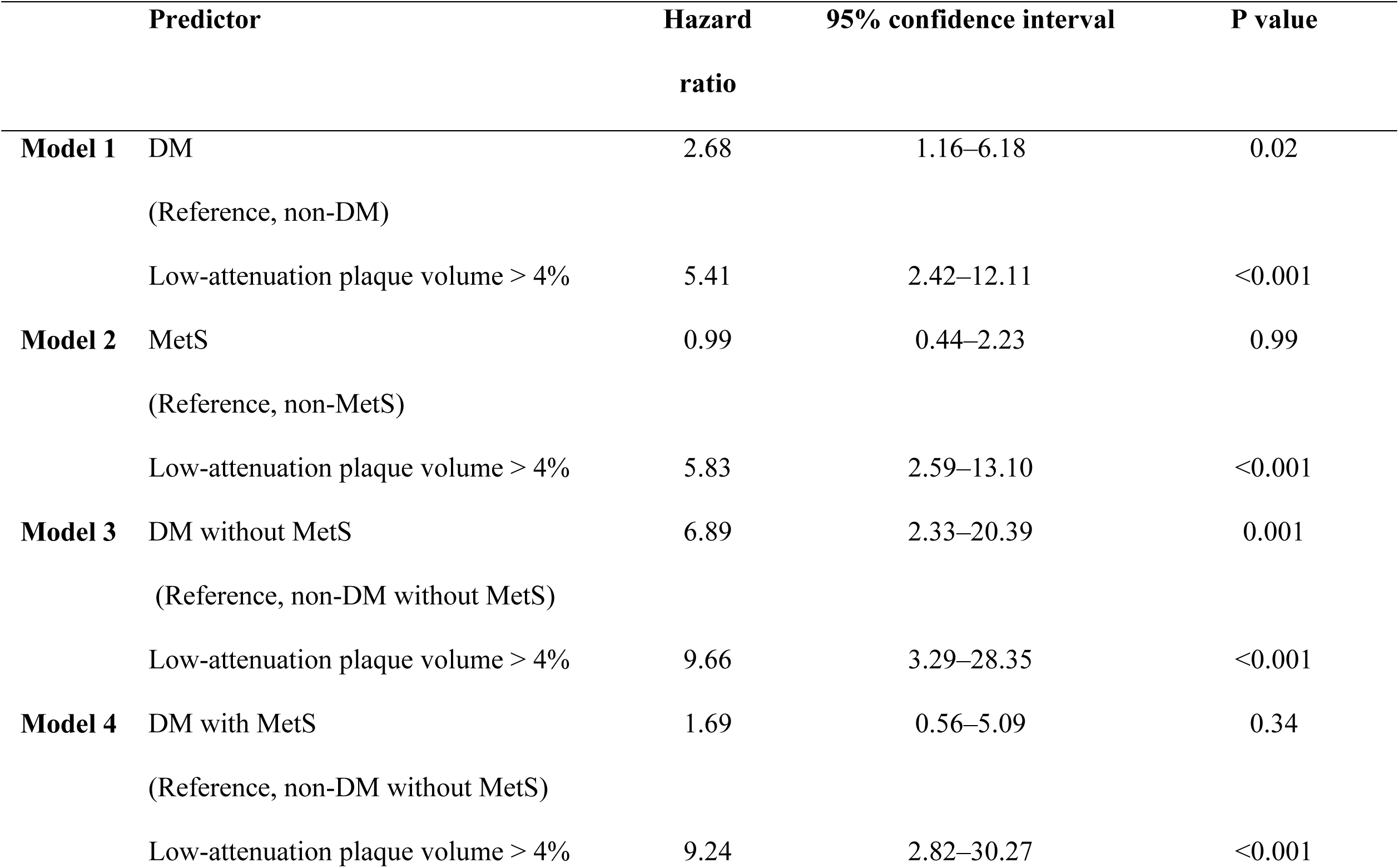
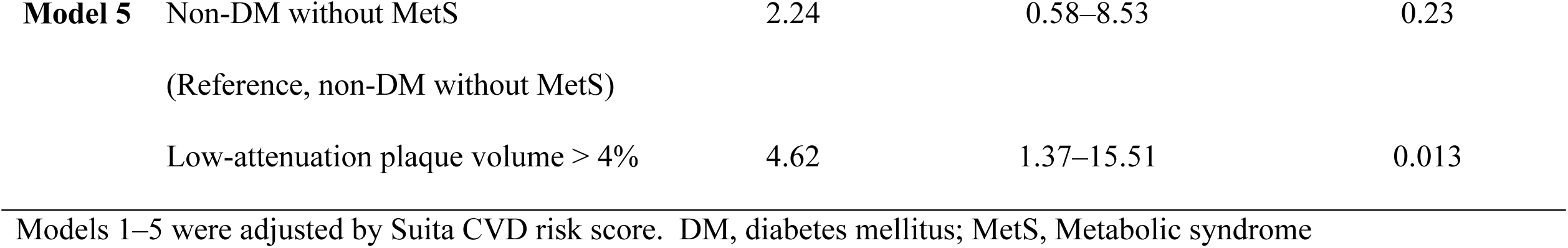
Cox proportional hazards analysis to predict major cardiovascular adverse events

## Discussion

This study investigated the association between distinct metabolic phenotypes according to the presence or absence of MetS and DM, and cardiovascular outcomes in symptomatic patients undergoing CCTA. The major findings of this study were as follows: (1) among the four groups, the worst prognosis was observed in non-MetS patients with DM compared to MetS patients with or without DM; (2) LAP volume > 4% was a robust predictor of MACE among the metabolic phenotypes; and (3) DM, independent of LAP volume > 4%, was a predictor of the primary endpoint; while MetS did not show a significant predictive value.

### Metabolic disorders, high-risk plaque burden, and outcomes

Several large clinical trials have demonstrated that myocardial ischemia is an important surrogate marker for improving outcomes in patients with stable CAD, whereas ischemia-guided management has a limited ability to prevent acute coronary events compared with optimized medical therapy ^20, 21^. These findings question the credibility of ischemia-guided management of patients with stable CAD, thus redirecting attention towards coronary microvascular dysfunction and high-risk plaque burden ^22–24^. An increased plaque burden, especially of non-calcified plaques, has been reported in patients with MetS ^25^. Yonetsu et al. used optical coherence tomography to demonstrate that MetS were associated with an increased burden of lipid-rich plaques ^25^. Although these findings indicate a potential link between obesity, metabolic disorders, and unfavorable coronary plaque features, there is limited knowledge regarding coronary plaque burden in distinct metabolic phenotypes with and without DM.

Previous clinical studies have reported an inverse association between BMI and cardiovascular prognosis (obesity paradox) ^7, 26, 27^. In patients with ASCVD and DM, Pagidipati et al. demonstrated that overweight or obese individuals had a lower cardiovascular risk than those with normal weight ^27^. In a sub-analysis of a large clinical CCTA trial of patients with chest pain, among distinct metabolic phenotypes, Kammerlander et al. demonstrated that metabolically unhealthy individuals without obesity exhibited an increased high-risk plaque burden and a high risk of ASCVD events ^7^. This paradoxically benign effect of obesity may be explained by its protective effect against atherosclerosis. Obese patients lack sarcopenia, have limited capacity for exercise, and have reduced mobility, all of which are associated with an increased incidence of cognitive decline, heart failure, and mortality ^28^. In addition, we observed increased fasting plasma glucose levels in patients with non-MetS DM. Hyperglycemia and insulin resistance have been reported to be key drivers of calcification in DM ^8, 11, 29^. Liu et al. showed that higher glucose levels and their variability are associated with plaque rupture in patients with ST-segment elevation myocardial infarction ^30^. These findings suggest that metabolic phenotypes can help identify patients at high risk of cardiovascular events in addition to a high-risk plaque burden.

### Epicardial adipose tissue, plaque characteristics, and outcomes

CAD is a chronic inflammatory disease with an underlying risk of metabolic disorders ^31^. A close relationship between abdominal visceral obesity and an increased coronary atherosclerotic burden has been reported ^4, 32^. Our results demonstrated that MetS patients with or without DM had increased EAT and LAP volumes, whereas non-MetS patients with DM had the worst outcomes, with lower EAT and LAP volumes, indicating an alternate pathophysiology of acute coronary syndromes in the latter. Our findings are consistent with the observations of Kammerlander et al., who demonstrated that both metabolically unhealthy obese and non-obese subjects exhibited increased high-risk plaques ^7^. The increased prevalence of obstructive CAD and vascular calcification (CP) in patients with DM offers a possible explanation for this finding^33^. Distinct plaque characteristics may reflect different (advanced or less advanced) stages of coronary atherosclerosis, resulting in different responses to lipid-lowering therapy ^34^. Furthermore, previous studies investigating plaque structural stress have demonstrated that microcalcifications contribute to increased stress, leading to plaque rupture and myocardial infarction ^35, 36^. These observations explain the poor outcomes of patients with DM without MetS in this study.

Although we found that the EAT volume was not correlated with outcomes, the mean CT value of the EAT was associated with cardiovascular outcomes (Table 3). EAT has been associated with coronary atherosclerosis, calcification, and cardiovascular outcomes, and has attracted attention as a therapeutic target ^18, 37–40^. This has motivated the development of imaging methods that enable the assessment of inflammation in the pericoronary adipose tissue, which interacts with the underlying vascular wall by producing proinflammatory adipokines ^41^. In a retrospective CCTA study, Oikonomou et al. demonstrated that a fat attenuation index > -70 HU around the epicardial coronary arteries predicts cardiovascular outcomes ^42^. Higher CT attenuation of the EAT indicates an increased inflammatory status, which supports our finding that non-MetS patients with DM have the worst prognosis.

In addition to plaque burden and metabolic disorders, coronary microvascular dysfunction may explain the association between non-MetS patients with DM and poorer outcomes ^43, 44^. In a systematic analysis, Kelshiker et al. demonstrated that patients with DM were more likely to have impaired coronary microvascular function, which presumably indicates a poor prognosis ^44^. Although the assessment of coronary microvascular dysfunction is appreciated, routine non-invasive evaluation methods are not available in daily clinical practice. Noninvasive assessment of the coronary plaque burden and metabolic phenotypes allows for further risk stratification of symptomatic patients undergoing CCTA.

### Study limitations

This study included a relatively small number of patients, and the event rates during follow-up were relatively low (< 5%). Our findings should be interpreted with caution since the DM with MET group was significantly older than the other groups with more obstructive CAD. Further studies are required to confirm these findings. Furthermore, although a unified definition is needed, the criteria for abdominal obesity vary among races ^45^. In the present study, we used quantitative VAT values obtained from CT scans to define abdominal obesity. Lastly, we did not perform laboratory tests such as HOMA-IR to measure insulin resistance, which is associated with inflammation ^46^ and plaque vulnerability ^8^. Instead, we calculated the TyG index and reported its significant association with the primary endpoint, independent of LAP volume > 4% (HR for TyG index, 1.78; 95% CI, 1.10–2.89; p = 0.019) (Model 1, in the supplementary Table 1).

## Conclusions

Individuals with DM alone have a significantly higher risk of developing MACE than those with MetS. This indicates that DM is an independent predictor of ASCVD events irrespective of the presence of obstructive CAD or high-risk plaque volume.

## Data Availability

Data is available from the corresponding author by reasonable request

## Acknowledgements

Not applicable

## Supplemental Material

Supplementary table 1

## Non-standard Abbreviations and Acronyms

ACS: acute coronary syndrome
ASCVD: Atherosclerotic cardiovascular disease
CACS: Coronary artery calcium score
CAD: coronary artery disease
CCTA: coronary computed tomography angiography
CP: Calcified plaques
DM: diabetes mellitus
LAP: Low-attenuation plaque
MACE: Major adverse cardiovascular events
MetS: Metabolic Syndrome
NCP: noncalcified plaque

